# Digital Access, Transportation, and Women’s Empowerment in Breast Cancer Screening Uptake Among Cambodian Women: Analysis of the Cambodia Demographic and Health Survey 2021-2022

**DOI:** 10.1101/2025.08.01.25332762

**Authors:** Samnang Um, Channnarong Phan, Daraden Vang, Tharuom Ny, Sothy Heng

## Abstract

The breast cancer cases are increasing, and it is the third leading cause of morbidity and mortality among cancer cases in women in Cambodia. This study explores how access to digital tools, media exposure, transportation, travel time to health facilities, and autonomy in health decisions relate to breast cancer screening among Cambodian women aged 15 to 49. After excluding 204 women who were unaware of breast or cervical cancer screening, the final weighted sample comprised 19,292 participants. The outcome was whether a woman had ever received a breast examination from a healthcare provider. We used multivariable logistic regression to assess associations between screening and key factors, adjusting for demographic and socioeconomic characteristics. Only 10.9% (95% CI: 9.7%–11.6%) of women had undergone a breast exam. Exposure to multiple forms of media was associated with a higher odd of screening (AOR = 1.47; 95% CI: 1.13–1.91). Phone ownership—whether a basic mobile phone (AOR = 1.35; 95% CI: 1.03–1.78) or smartphone (AOR = 1.37; 95% CI: 1.03–1.82)—was also positively associated. In contrast, longer travel times of over 30 minutes (AOR = 0.55; 95% CI: 0.39–0.78) and a lack of autonomy in healthcare decisions (AOR = 0.70; 95% CI: 0.52–0.94) were associated with reduced screening. Wealthier women had greater odds of being screened (AOR = 1.86; 95% CI: 1.40–2.48). The findings underscore the need to strengthen communication, improve access to care, and support women’s decision-making to increase screening rates in Cambodia.

## Introduction

In 2022, the Global Cancer Observatory estimated approximately 24% of new cases (2,296,840 new breast cancer cases), out of a total of 9,664,889 cancer cases reported in 2022, and 666,103 (or 15.4%) deaths due to breast cancer globally, represented a significant public health burden (1). The rising burden is particularly in low- and middle-income countries (LMICs), including Cambodia, where late-stage diagnosis, limited access to screening, and underdeveloped cancer control infrastructure contribute to poorer outcomes (2-4).

In Cambodia, breast cancer accounts for a significantly increased rate of breast cancer morbidity and mortality, accounting for approximately 20% of new cases (2,116 new female cancer cases), out of a total of 10,624 cancer cases reported in 2022, and 917 (or 14%) deaths due to breast cancer (5). Although national incidence data remain limited, hospital-based registries and regional studies have reported an increasing trend in breast cancer cases (3, 5-10). Early detection through breast cancer screening, especially clinical breast examination (CBE), is a cost-effective and feasible method in resource-limited settings and has been shown to improve survival rates (4, 11-19). Nonetheless, screening coverage remains low in Cambodia, with only 11% of women aged 15–49 reporting that they had ever checked for breast cancer with physicians or other healthcare professionals, with persistent socioeconomic, cultural, and geographic barriers impeding access (3-5,7). Moreover, previous studies indicated that knowledge about breast self-examination (BSE) in Cambodia was limited, with approximately 60% of Cambodian women being unaware of BSE (3). To address this, strengthening cancer screening in Cambodia requires ongoing efforts. The Cambodian Ministry of Health has prioritized it in the National Action Plan for Cervical Cancer Prevention and Control (2019-2023) and the National Cancer Control Plan (2025-2030), as noted in (13, 17).

Recent technological developments offer new opportunities to enhance awareness and uptake of screening services. Mobile phone ownership is high among Cambodian women, exceeding 80%, and internet penetration is expanding rapidly [6]. Media exposure—through television, radio, and newspapers—remains a powerful channel for public health messaging and outreach interventions within the community, especially in LMICs [7,8]. Studies from other settings have shown that women with digital access and frequent media exposure can improve their cancer knowledge and promote screening behaviors [9,10]. However, these relationships have not been comprehensively studied in the Cambodian context.

In addition to communication technologies, access to transportation plays a key role in healthcare utilization. In Cambodia, motorcycles are the most common form of personal transport, particularly in rural areas [11]. Motorcycle ownership may enable women to overcome geographic barriers by reducing travel time and increasing independence in reaching health services [12]. Yet, its specific role in facilitating breast cancer screening has not been systematically examined.

Barriers to healthcare access—such as distance to facilities, financial constraints, permission from others, or unwillingness to seek care alone—also remain prevalent [13]. Women’s autonomy in healthcare decision-making is another critical factor that may influence the uptake of preventive services like cancer screening, but it remains understudied in Cambodia.

A recent study using CDHS 2021–2022 data examined the socio-demographic and behavioral determinants of breast and cervical cancer screening in Cambodian women aged 15–49 years, focusing on factors such as women’s age, marital status, education, wealth index, geographical regions, place of residence, health insurance coverage, health facility visits within the past year, number of children ever born, smoking, alcohol consumption, contraceptive use, and body mass index [14]. While that analysis provided foundational insights, it did not explore the influence of emerging digital access, transportation, travel time to the facility, and women’s decision-making autonomy [14].

This study addresses these knowledge gaps by examining how media access, smartphone ownership, internet use, motorcycle ownership, travel time to the facility, and women’s decision-making autonomy are associated with breast cancer screening among Cambodian women of reproductive age after adjusting for socio-demographics. Using nationally representative data analysis, the findings would inform more effective and equitable interventions to increase screening coverage and reduce breast cancer burden in Cambodia.

## Methods

### Ethical statement

The Cambodia National Ethics Committee for Human Health Research (NECHR) approved the data collection tools and procedures for CDHS 2021-2022 for Health Research on May 10, 2021 (Ref # 83 NECHR), and ICF’s Institutional Review Board (IRB) in Rockville, Maryland, USA. Written informed consent was obtained from all participants prior to data collection. For respondents under 18 years of age, consent was obtained from a parent or guardian. This study used de-identified secondary data and was therefore exempt from additional institutional ethical approval.

### Data Source

This study utilized data from the Cambodia Demographic and Health Survey (CDHS) 2021–2022, a nationally representative cross-sectional survey designed to provide estimates of key demographics, reproductive health, and nutrition indicators for women and children. Data was collected from September 15, 2021, to February 15, 2022. The CDHS employed a two-stage stratified cluster sampling method, selecting women from all provinces of Cambodia. In the initial stage, 709 enumeration areas (EAs) (241 urban areas and 468 rural areas) were selected. In the second stage, an equal systematic sample of 25–30 households was selected from each cluster of 21,270 families. In total, 19,496 women aged 15–49 were interviewed face-to-face, using the survey questionnaire, with a response rate of 98.2%. The final CDHS 2021-2022 report details have been published (8).

### Study Population

The study population consisted of women aged 15–49 years who responded to questions about breast cancer screening examined by a health professional, yielding a total of 19,292 weighted responses. Women with missing data on the primary outcome or key exposure variables were excluded from the analysis (n = 204). Overall, 204 women reported that they did not know about breast and cervical cancer screening, and were excluded. Data restriction resulted in a final analysis with 19,292 weighted samples of women.

## Measurement Variables

### Outcome Variable

The outcome was breast cancer screening uptake, measured by asking women if they had ever had their breasts examined by a doctor or other health care provider to check for cancer. This examination encompassed both clinical breast examinations (CBEs), where providers manually palpate for lumps or changes, and imaging techniques, such as mammograms (8). This variable was coded as binary (1 = Yes, 0 = No) (8).

### Independent Variables

Digital Access consisted of type of mobile phone ownership (no phone, non-smartphone, and smartphone), internet use within the past 12 months (yes vs no), and media exposure measured by the frequency of reading newspapers, listening to the radio, and watching television, then was categorized into (none, exposure to one source, or exposure to two or more sources) (8).

Transportation access was measured by whether the household owned a motorcycle (yes vs. no). Additional measures of healthcare access included the time required to travel to the nearest health facility, categorized as near (≤10 minutes), moderate (11–30 minutes), or far (>30 minutes).

Women’s autonomy in healthcare decision-making was categorized into three groups based on who makes decisions regarding the respondent’s healthcare: autonomous (respondent alone), joint (shared decision-making), and non-autonomous (decisions made by others).

Covariate variables included women’s age (15–29, 30–39, and 40–49 years), education level (no education, incomplete primary, complete primary, incomplete secondary, complete secondary and higher), and household wealth status (classified into quintiles from poorest to richest) (8). Additional geographic variables included place of residence (urban or rural) (8).

### Statistical Analysis

All analyses accounted for the complex survey design, utilizing sampling weights, clustering, and stratification as provided by the CDHS—descriptive statistics summarized sample characteristics and breast cancer screening coverage, reported as weighted counts and percentages.

Bivariate associations were tested using chi-square tests and unadjusted logistic regression models. Multivariable logistic regression with survey design adjustment was used to examine factors associated with breast cancer screening. The analysis accounted for the complex survey design, including stratification, clustering, and sampling weights using the svy: prefix in STATA. The following independent variables were included: media exposure, smartphone ownership, internet use in the last 12 months, motorcycle ownership, time to reach health care, decision-making autonomy, current age group, household wealth index, education level, occupation, and place of residence. The final results of the multivariable binary logistic regression were reported as adjusted odds ratios (AOR) and 95% confidence intervals (CIs), along with the p-value. Statistical significance was defined as p < 0.05.

The goodness-of-fit was assessed using the Hosmer-Lemeshow test with ten groups. Predicted probabilities were generated post-estimation to evaluate model discrimination via the area under the receiver operating characteristic (ROC) curve. Statistical significance was determined at a p-value <0.05.

Multicollinearity among the independent variables was assessed using Variance Inflation Factors (VIFs) derived from a linear regression model that included all predictors. A VIF value greater than 5 was considered indicative of problematic multicollinearity. Variables included in the multicollinearity assessment were media exposure, smartphone ownership, internet use, motorcycle ownership, decision-making group, women’s age, wealth index, education, occupation, and place of residence. The VIF values for all predictors ranged from 1.01 to 1.79, with a mean VIF of 1.33, indicating low multicollinearity among the independent variables. No variable exceeded the threshold of 5, suggesting that multicollinearity was not a concern in the model.

We assessed potential effect modification between smartphone ownership and age by including an interaction term in the multivariable logistic regression model using the factor-variable notation. The significance of interaction terms was evaluated using adjusted Wald tests with the testparm command under the complex survey design.

## Results

### Description of the study samples

Among 19,292 weighted women aged 15–49 years, the average age was 31.0 years (95% CI: 30.9– 31.2). The majority had no mass media exposure (71.9%), owned a smartphone (77.5%), and had used the internet in the past year (63.6%). Over half (60.8%) owned a motorcycle, and half (50.8%) lived within a 10-minute distance of a health facility. About 44% reported autonomy in healthcare decision-making, with 23.1% in the richest and 17.4% in the poorest group. Most women had incomplete secondary education (35.6%) or incomplete primary education (29.3%). Approximately 42.4% worked in informal jobs, while 30.4% held professional roles. A larger proportion lived in urban areas (57.5%). Overall, only 10.9% (95% CI: 9.7%-11.6%) of women reported ever having a breast exam by health providers (**Table 1)**.

**Table 1.** Distribution of Characteristics of Media Access, Digital Access, Motorcycle Access, Healthcare decision-making, and Socioeconomic Status Associated with Breast Exam Uptake Among Women Aged 15–49 in Cambodia, CDHS 2021–2022 (N=19,292 weighted).

| Variable | Total | Breast Exam Uptake |  | P-value |
| --- | --- | --- | --- | --- |
|  |  | Yes | No |  |
|  | N (%) | n (%) | n (%) |  |
| Media Access |  |  |  |  |
| None | 13,865 (71.9) | 1,321 (9.5) | 12,544 (90.5) | <0.001 |
| One source | 4,433 (23.0) | 564 (12.7) | 3,869 (87.3) |  |
| Two or more sources | 993 (5.1) | 161 (16.2) | 832 (83.8) |  |
| Smartphone Ownership |  |  |  |  |
| No phone | 2,917 (15.1) | 210 (7.2) | 2,707 (92.8) | <0.001 |
| Non-smartphone | 1,423 (7.4) | 156 (11.0) | 1,266 (89.0) |  |
| Smartphone | 14,952 (77.5) | 1,679 (11.2) | 13,273 (88.8) |  |
| Internet use (last 12 months) |  |  |  |  |
|  |  | Yes | No |  |
|  | N (%) | n (%) | n (%) |  |
| No | 7,020 (36.4) | 661 (9.4) | 6,359 (90.6) | 0.016 |
| Yes | 12,272 (63.6) | 1,385 (11.3) | 10,887 (88.7) |  |
| Motorcycle Ownership |  |  |  |  |
| No | 7,570 (39.2) | 932 (12.3) | 6,638 (87.7) | <0.001 |
| Yes | 11,721 (60.8) | 1,113 (9.5) | 10,608 (90.5) |  |
| Travel time to the facility |  |  |  |  |
| ≤10 minutes | 9,811 (50.8) | 1,208 (12.3) | 8,602 (87.7) | <0.001 |
| 11–30 minutes | 8,397 (43.5) | 783 (9.3) | 7,614 (90.7) |  |
| >30 minutes | 1,084 (5.6) | 54 (5.0) | 1,030 (95.0) |  |
| Healthcare decision-making |  |  |  |  |
| Autonomous | 5,866 (30.4) | 4,983 (84.9) | 883 (15.1) | <0.001 |
| Joint | 6,379 (33.1) | 5,619 (88.1) | 760 (11.9) |  |
| Not autonomous | 7,043 (36.5) | 6,641 (94.3) | 402 (5.7) |  |
| Women's mean age in years (95% CI) | 31.0 (30.9–31.2) | 35.2 (34.6–35.7) | 30.6 (30.4–30.7) |  |
| 15-29 | 8,444 (43.8) | 464 (5.5) | 7,980 (94.5) | <0.001 |
| 30-39 | 6,589 (34.2) | 967 (14.7) | 5,622 (85.3) |  |
| 40-49 | 4,258 (22.1) | 614 (14.4) | 3,644 (85.6) |  |
| Wealth index |  |  |  |  |
| Poorest | 3,355 (17.4) | 246 (7.3) | 3,108 (92.7) | <0.001 |
| Poorer | 3,451 (17.9) | 262 (7.6) | 3,189 (92.4) |  |
| Middle | 3,790 (19.6) | 314 (8.3) | 3,476 (91.7) |  |
| Richer | 4,243 (22.0) | 434 (10.2) | 3,808 (89.8) |  |
| Richest | 4,453 (23.1) | 789 (17.7) | 3,664 (82.3) |  |
| Education level |  |  |  |  |
| No education | 2,232 (11.6) | 197 (8.8) | 2,035 (91.2) | 0.0005 |
| Incomplete primary | 5,652 (29.3) | 608 (10.8) | 5,044 (89.2) |  |
| Complete primary | 1,823 (9.4) | 204 (11.2) | 1,619 (88.8) |  |
| Incomplete secondary | 6,874 (35.6) | 654 (9.5) | 6,220 (90.5) |  |
| Complete secondary | 1,320 (6.8) | 168 (12.7) | 1,153 (87.3) |  |
|  |  | Yes | No |  |
|  | N (%) | n (%) | n (%) |  |
| Higher | 1,390 (7.2) | 215 (15.5) | 1,175 (84.5) |  |
| **Occupation** |  |  |  |  |
| Not working | 5,249 (27.2) | 427 (8.1) | 4,823 (91.9) | <0.001 |
| Professional | 5,866 (30.4) | 896 (15.3) | 4,970 (84.7) |  |
| Informal | 8,176 (42.4) | 723 (8.8) | 7,453 (91.2) |  |
| **Place of residence** |  |  |  |  |
| Rural | 8,205 (42.5) | 1,075 (13.1) | 7,131 (86.9) | <0.001 |
| Urban | 11,086 (57.5) | 971 (8.8) | 10,115 (91.2) |  |

Survey weights are applied to obtain weighted percentages.

**Table 2.**
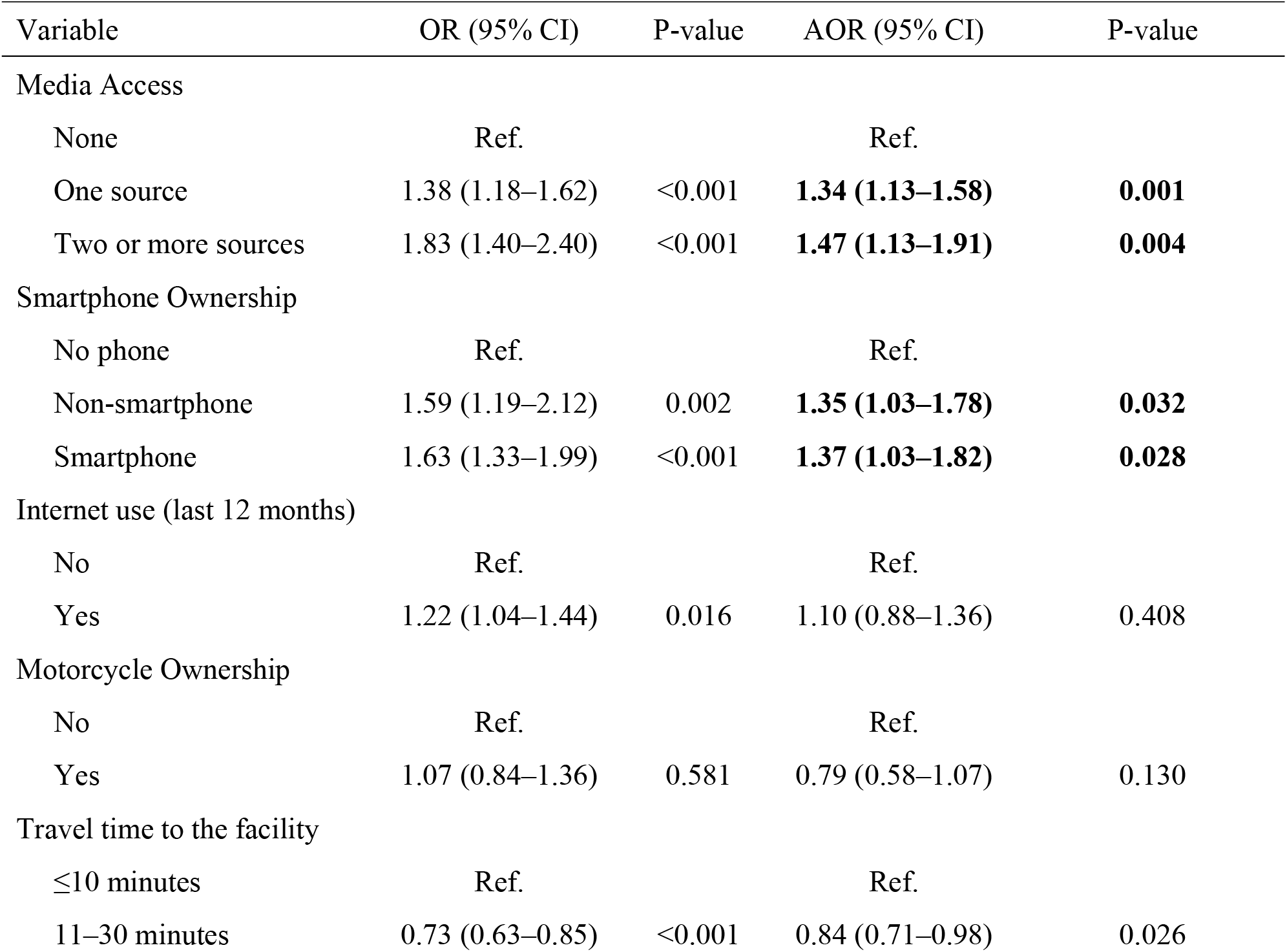
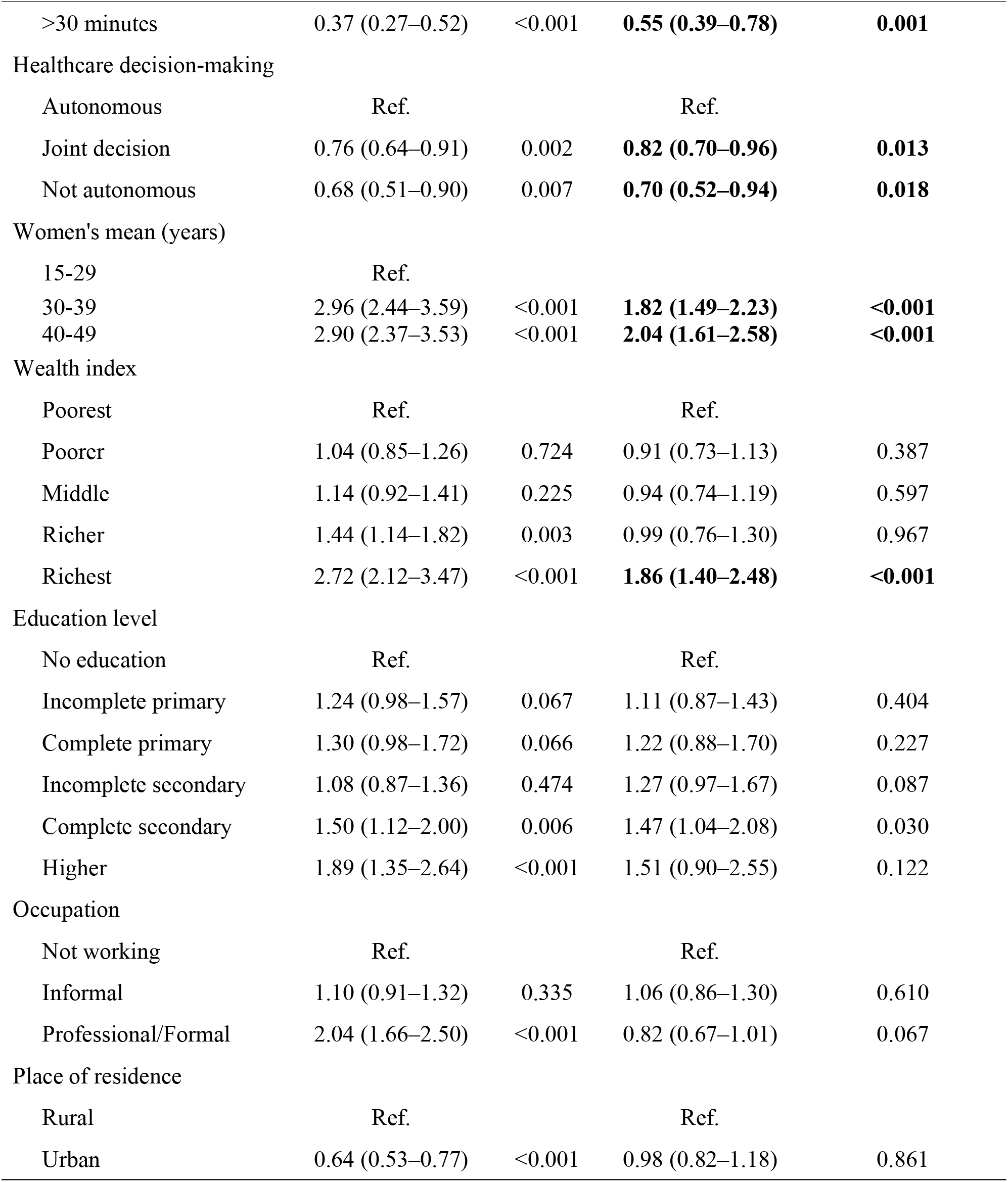
Unadjusted and adjusted logistic regression analysis of factors associated with breast exam uptake among women aged 15–49 years, CDHS 2021–2022 (N=19,287)

| Variable | OR (95% CI) | P-value | AOR (95% CI) | P-value |
| --- | --- | --- | --- | --- |
| Media Access |  |  |  |  |
| None | Ref. |  | Ref. |  |
| One source | 1.38 (1.18–1.62) | <0.001 | **1.34 (1.13–1.58)** | **0.001** |
| Two or more sources | 1.83 (1.40–2.40) | <0.001 | **1.47 (1.13–1.91)** | **0.004** |
| Smartphone Ownership |  |  |  |  |
| No phone | Ref. |  | Ref. |  |
| Non-smartphone | 1.59 (1.19–2.12) | 0.002 | **1.35 (1.03–1.78)** | **0.032** |
| Smartphone | 1.63 (1.33–1.99) | <0.001 | **1.37 (1.03–1.82)** | **0.028** |
| Internet use (last 12 months) |  |  |  |  |
| No | Ref. |  | Ref. |  |
| Yes | 1.22 (1.04–1.44) | 0.016 | 1.10 (0.88–1.36) | 0.408 |
| Motorcycle Ownership |  |  |  |  |
| No | Ref. |  | Ref. |  |
| Yes | 1.07 (0.84–1.36) | 0.581 | 0.79 (0.58–1.07) | 0.130 |
| Travel time to the facility |  |  |  |  |
| ≤10 minutes | Ref. |  | Ref. |  |
| 11–30 minutes | 0.73 (0.63–0.85) | <0.001 | 0.84 (0.71–0.98) | 0.026 |
| >30 minutes | 0.37 (0.27–0.52) | <0.001 | **0.55 (0.39–0.78)** | **0.001** |
| Healthcare decision-making |  |  |  |  |
| Autonomous | Ref. |  | Ref. |  |
| Joint decision | 0.76 (0.64–0.91) | 0.002 | **0.82 (0.70–0.96)** | **0.013** |
| Not autonomous | 0.68 (0.51–0.90) | 0.007 | **0.70 (0.52–0.94)** | **0.018** |
| Women's mean (years) |  |  |  |  |
| 15-29 | Ref. |  |  |  |
| 30-39 | 2.96 (2.44–3.59) | <0.001 | **1.82 (1.49–2.23)** | **<0.001** |
| 40-49 | 2.90 (2.37–3.53) | <0.001 | **2.04 (1.61–2.58)** | **<0.001** |
| Wealth index |  |  |  |  |
| Poorest | Ref. |  | Ref. |  |
| Poorer | 1.04 (0.85–1.26) | 0.724 | 0.91 (0.73–1.13) | 0.387 |
| Middle | 1.14 (0.92–1.41) | 0.225 | 0.94 (0.74–1.19) | 0.597 |
| Richer | 1.44 (1.14–1.82) | 0.003 | 0.99 (0.76–1.30) | 0.967 |
| Richest | 2.72 (2.12–3.47) | <0.001 | **1.86 (1.40–2.48)** | **<0.001** |
| Education level |  |  |  |  |
| No education | Ref. |  | Ref. |  |
| Incomplete primary | 1.24 (0.98–1.57) | 0.067 | 1.11 (0.87–1.43) | 0.404 |
| Complete primary | 1.30 (0.98–1.72) | 0.066 | 1.22 (0.88–1.70) | 0.227 |
| Incomplete secondary | 1.08 (0.87–1.36) | 0.474 | 1.27 (0.97–1.67) | 0.087 |
| Complete secondary | 1.50 (1.12–2.00) | 0.006 | 1.47 (1.04–2.08) | 0.030 |
| Higher | 1.89 (1.35–2.64) | <0.001 | 1.51 (0.90–2.55) | 0.122 |
| Occupation |  |  |  |  |
| Not working | Ref. |  | Ref. |  |
| Informal | 1.10 (0.91–1.32) | 0.335 | 1.06 (0.86–1.30) | 0.610 |
| Professional/Formal | 2.04 (1.66–2.50) | <0.001 | 0.82 (0.67–1.01) | 0.067 |
| Place of residence |  |  |  |  |
| Rural | Ref. |  | Ref. |  |
| Urban | 0.64 (0.53–0.77) | <0.001 | 0.98 (0.82–1.18) | 0.861 |

### Association with breast cancer screening in Chi-Square

The prevalence of breast cancer screening (ever having a breast exam by a health professional) was 10.6% (95% CI: 9.7%–11.6%). Women who had undergone a breast exam were, on average, older, with a mean age of 35.2 years (95% CI: 34.6–35.7), compared to 30.6 years (95% CI: 30.4– 30.7) among those who had not. Screening prevalence increased with age: 5.5% among women aged 15–29 years, 14.7% among those aged 30–39 years, and 14.4% among women aged 40–49 years. These differences in breast exam uptake across age groups were statistically significant (p < 0.001). Women’s exposure to mass media was significantly associated with breast exam uptake (p < 0.001). The proportion of women who had ever undergone a breast exam increased from 9.5% among those with no media exposure to 16.2% among those exposed to two or more sources. Smartphone ownership and recent internet use were also associated with higher uptake (p < 0.001 and p = 0.016, respectively). Specifically, 11.2% of smartphone users had ever undergone a breast exam, compared to 7.2% of women who did not own a smartphone. Access to healthcare services was a crucial factor in this decision. Uptake was highest among women living within 10 minutes of a health facility (12.3%) and lowest among those living more than 30 minutes away (5.0%) (p < 0.001). Similarly, women with autonomy in healthcare decision-making had a higher uptake (15.1%) compared to those without (10.7%) (p = 0.0015). Breast exam uptake increased with household wealth, ranging from 7.3% in the poorest quintile to 17.7% in the richest (p < 0.001). A similar pattern was seen by education level; women with higher education had greater uptake (15.5%) compared to those with no education (8.8%) (p = 0.0005). Women in professional occupations were more likely to have had a breast exam (15.3%) than those not working (8.1%) or working in informal jobs (8.8%) (p < 0.001). Notably, rural women reported higher uptake (13.1%) than urban women (8.8%) (p < 0.001), contrary to typical patterns in healthcare access **(Table 1**).

### Factors Associated with Breast Exam Uptake in Multivariable Analysis

In the final multivariable binary logistic regression model was statistically significant (F(24, 637) = 8.71, p < 0.001), indicating that the set of predictors explained a substantial amount of variation in breast cancer screening.

In the fully adjusted model, several factors remained significantly associated with breast exam uptake among women aged 15–49 years. Women exposed to mass media had higher odds of breast exam uptake compared to those with no media exposure; those with one source had 1.34 times the odds (AOR=1.34; 95% CI: 1.13–1.58), and those with two or more sources had 1.47 times the odds (AOR = 1.47; 95% CI: 1.13–1.91). Similarly, phone ownership was positively associated with screening uptake. Both non-smartphone users (AOR = 1.35; 95% CI: 1.03–1.78) and smartphone users (AOR = 1.37; 95% CI: 1.03–1.82) had significantly higher odds of breast exam uptake compared to women without phones. Longer travel time to the nearest health facility was inversely related to breast exam uptake: women living 11–30 minutes away had 16% lower odds (AOR= 0.84, 95% CI: 0.71–0.98), and those living more than 30 minutes away had 45% lower odds (AOR=0.55, 95% CI: 0.39–0.78), compared to women living within 10 minutes. Healthcare decision-making autonomy was also significant; women who made decisions jointly with others (AOR = 0.82, 95% CI: 0.70–0.96) or were not autonomous (AOR = 0.70, 95% CI: 0.52–0.94) had significantly lower odds of screening compared to women who made healthcare decisions autonomously. Age remained a strong predictor of screening uptake. Women aged 30–39 years had 1.8 times the odds (AOR = 1.8; 95% CI: 1.49–2.23), and those aged 40–49 years had twice the odds (AOR = 2.04, 95% CI: 1.61–2.58) of undergoing a breast exam compared to women aged 15–29 years. Regarding socioeconomic status, only women in the richest wealth quintile had significantly higher odds of screening (AOR = 1.86, 95% CI: 1.40–2.48) compared to those in the poorest quintile. Educational attainment was associated with uptake only at the level of completing secondary school (AOR = 1.47, 95% CI: 1.04–2.08). Occupation and urban versus rural residence were not statistically significant after adjustment for other factors. Internet use and motorcycle ownership did not show significant associations in the adjusted model.

The Hosmer-Lemeshow goodness-of-fit test indicated adequate model fit (F(9,652) = 0.42, p = 0.925). The ROC curve analysis yielded an area under the curve (AUC) of 0.653 (95% CI: 0.638– 0.668), indicating moderate discriminative ability.

The interaction between smartphone ownership and age was statistically significant (Adjusted Wald test: F(8, 653) = 9.54, p < 0.001), indicating that the association between smartphone ownership and breast cancer screening varied across age groups. For example, among women aged 40–49, smartphone users had significantly higher odds of screening (AOR = 2.22; 95% CI: 1.34–3.68; p = 0.002), compared to non-users aged 15–29. Similarly, among those aged 30–39, smartphone use was associated with increased screening (AOR = 1.71; 95% CI: 1.07–2.75; p = 0.026). However, no significant effect was observed among the youngest age group (15–29 years; AOR = 0.89; 95% CI: 0.54–1.46; p = 0.650) (**S1 Table**). In contrast, the interaction between media exposure and smartphone ownership was not statistically significant (F(4, 657) = 0.63, p = 0.642), suggesting that their effects on breast cancer screening were independent.

## Discussion

This study highlights several important determinants of breast exam uptake among women aged 15–49 in Cambodia. Consistent with previous research from LMICs, media exposure was strongly associated with higher screening rates, with women exposed to two or more media sources having 47% higher odds of having had a breast exam (AOR = 1.47, 95% CI: 1.13–1.91). Mass media campaigns in Cambodia, including radio and television programs supported by the Ministry of Health and partners, likely play a key role in increasing awareness and encouraging early detection (13, 17).

The finding that owning a non-smartphone was significantly associated with increased odds of undergoing a breast exam (AOR = 1.35; 95% CI: 1.03–1.78; p = 0.032) suggests that even basic mobile phone access may facilitate communication about health services in Cambodia. Interestingly, smartphone ownership also showed a significant positive association (AOR = 1.37; 95% CI: 1.03–1.82; p = 0.028). This suggests that while both basic phones and smartphones play a role in supporting outreach through voice calls and SMS reminders, smartphones, with their greater potential for accessing health-related information, receiving appointment reminders, or engaging with digital health content, demonstrate a stronger, more statistically significant association with screening uptake. However, despite this statistical strength, the magnitude of the association (AOR) is similar to that of non-smartphones. This similarity in AORs, despite the difference in p-values, may reflect underlying challenges in the Cambodian context, such as limited digital literacy among women, unequal access to relevant health apps, and a lack of widely adopted or tailored digital health campaigns that could fully leverage smartphone capabilities for preventive health behaviors (20, 21). These findings align with regional evidence from Southeast Asia, where mobile phone access has been shown to enhance uptake of maternal and reproductive health services in resource-limited settings (22-26). Expanding mobile health (mHealth) initiatives that are inclusive of both smartphone and non-smartphone users—particularly through voice and text-based interventions, as well as increasingly sophisticated yet user-friendly digital health content—could strengthen outreach and improve breast cancer screening coverage, especially among underserved populations in Cambodia (27). Future research should explore not only device ownership but also women’s digital literacy, usage patterns, and engagement with health-related content to understand better and leverage the role of mobile technology in preventive care.

Longer travel time to health facilities was strongly linked to lower breast exam uptake: women living more than 30 minutes away had 46% lower odds compared to those within 10 minutes (AOR = 0.54, 95% CI: 0.38–0.76, p = 0.001). This supports the emphasis of Cambodia’s National Strategic Plan for Cancer Control on decentralizing screening services to reduce travel burdens (12, 13, 17). Despite progress, physical distance and transportation remain barriers, especially for poorer women. Women’s autonomy in healthcare decision-making was positively associated with screening; those making joint decisions had 19% lower odds (AOR = 0.81, 95% CI: 0.69–0.94, p = 0.007), and those not autonomous had 31% lower odds (AOR = 0.69, 95% CI: 0.51–0.94, p = 0.017) compared to autonomous women. The result aligns with regional evidence that empowerment facilitates the use of preventive care (28-30). Cambodia’s community health worker programs aim to enhance women’s agency but may require scaling up. The wealthiest women had more than twice the odds of undergoing a breast exam compared to the poorest (AOR = 2.13, 95% CI: 1.60–2.84, p < 0.001), reflecting persistent socioeconomic disparities despite health equity initiatives, such as the Health Equity Fund (7, 31-36). Education level and urban residence were not significantly associated after adjustment, indicating that access and empowerment may be stronger determinants in Cambodia’s context. These findings reinforce the need to strengthen media campaigns, improve healthcare access, and empower women to reduce breast cancer screening inequalities in Cambodia (17, 33).

## Policy Implications

To address the persistently low rates of breast cancer screening among Cambodian women, especially those in rural and lower-income communities, a multifaceted approach that integrates digital health solutions, community-based outreach, and strengthened social protection mechanisms is essential. Evidence from Cambodia’s implementation of the Health Equity Fund (HEF) suggests that removing financial barriers through targeted subsidies significantly increases service utilization among the poorest households, including maternal and reproductive health services (37-39). Moreover, leveraging the role of Village Health Support Groups (VHSGs) and community health workers has proven effective in promoting preventive health behaviors and empowering women with knowledge and agency (33, 40). Integrating these strategies with digital innovations—such as mobile health messaging and appointment reminders—could further enhance access and adherence to breast cancer screening services, particularly among digitally connected women in underserved areas.

## Limitations

This study has several limitations. The cross-sectional design limits the ability to infer causal relationships between determinants and breast exam uptake. Data on breast exam history were self-reported, which may introduce recall bias or social desirability bias. The survey did not differentiate between clinical breast exams performed by health professionals and self-examinations, which limits specificity in interpreting the screening behaviors. Additionally, essential factors such as cultural beliefs, detailed knowledge about breast cancer, and quality of health services were not captured, potentially leading to residual confounding. Lastly, geographic accessibility was measured by travel time, but it did not account for other access barriers, such as transportation costs or facility readiness.

## Conclusion

Breast exam uptake among Cambodian women is influenced by factors such as media exposure, socioeconomic status, access to healthcare, and women’s autonomy in decision-making. To improve early breast cancer detection, health programs should enhance multi-channel media campaigns and expand decentralized screening services to reduce geographic barriers. Empowering women to participate actively in health decisions is also critical. Targeted strategies are needed to address socioeconomic disparities, ensuring vulnerable populations benefit equally. Future research should explore knowledge, attitudes, and health system factors to inform comprehensive breast cancer screening policies in Cambodia.

## Data Availability

This study used the 2021-2022 Cambodia Demographic and Health Survey (CDHS) datasets. The DHS data are publicly available from the website at (URL:https://www.dhsprogram.com/data/available-datasets.cfm).

## Acknowledgments

We thank the Ministry of Planning and the Ministry of Health of Cambodia for granting access to the CDHS dataset. We also acknowledge ICF International for technical support in survey implementation.

## Abbreviations

ACS: American Cancer Society
AOR: Adjusted odds ratio
BMI: Body mass index
CDHS: Cambodia Demographic Health Survey
EA: Enumeration areas
HPV: Human papillomavirus
NCDs: Noncommunicable diseases
PPS: Probability proportional to size
VIA: Visual inspection with acetic acid
WRA: Women at reproductive age
WHO: World Health Organization.

## Supporting information

**S1 Table**. Adjusted Odds Ratios (AOR) for Breast Cancer Screening by Smartphone Ownership and Age Group (Interaction Model)

